# Personalized prophylactic antiemetic regimens for the control of chemotherapy-induced nausea and vomiting by pharmacogenetic analysis of three receptors genes: *HTR3A, HTR3B, TACR1*

**DOI:** 10.1101/2024.12.29.24319754

**Authors:** Winnie Yeo, Frankie KF Mo, JingHan Huang, Horatio L. Yeo, Nicholas W.H. Ko, Leung V. Li, Thomas K.H. Lau, Kwai T. Lai, Elizabeth Pang, Menglin Ou, Suk-Ling Ma, Nelson L.S. Tang

## Abstract

**Background:** Contemporary prophylactic antiemetic regimens have improved the control of chemotherapy-induced nausea and vomiting (CINV). However, many patents still have suboptimal control with over 50% still suffering from nausea. We postulate that an individual’s pharmacogenetic profile may aid in optimizing the use of antiemetic prophylaxis. This study aimed to correlate the genetic determinants of individual patients with the efficacy of the prophylactic antiemetic regimens each received.

**Methods:** Breast cancer patients who were enrolled in 2 previously reported prospective antiemetic studies consented for the present pharmacogenetic study. Prior to highly emetogenic doxorubicin and cyclophosphamide (AC) (neo)adjuvant chemotherapy, they received a combination of antiemetic prophylaxis: regimen A and B were respectively aprepitant/ondansetron/dexamethasone with or without olanzapine; regimen C was netupitant/palonosetron/dexamethasone. Effectiveness of antiemetic regimens were mainly assessed by complete protection rate (CP) rates. Patients’ genotypes in 3 genes, HTR3A, HTR3B and TACR1, were analyzed.

**Findings:** Homozygous TT (p.129Tyr) genotype of a non-nonsynonymous variant (rs1176744) in HTR3B and homozygous GG of rs3821313 genotype in TACR1 had better outcome with regimen B (when olanzapine was combined with aprepitant/ondansetron/dexamethasone). Digenic interaction analysis further reveals interaction between rs1176744 (HTR3B) and rs3821313 (TACR1). Patients who were both homozygote T of rs1176744 and homozygote G of rs3821313 achieved the highest CP rate with regimen B (10/12 patients; 83%), in contrast to only 29% (7/24 of patients) given regimen A (p= 0.0027). Patients who were homozygote for G alleles in both rs1176722 of HRB3A and rs3821313 of TACR1 showed the poorest response to regimen A with CP rate of 17% (2/12), while patients given regimen B had the highest CP rate of 70% (7/10) (p= 0.0159). The findings were confirmed upon logistic regression adjusted for clinical factors.

**Interpretation:** The present study confirmed our hypothesis that among Chinese breast cancer patients who received AC, the selection of optimal antiemetic prophylaxis may be aided by assessing an individual’s pharmacogenetic profile. It also highlights a new phenomenon of digenic interaction that has not been known before for pharmacogenetic analysis.

**Funding:** This study was supported by an education grant from Madam Diana Hon Fun Kong Donation for Cancer Research.

**Research in context:** *Evidence before this study:* Although contemporary antiemetic prophylaxis has improved control of chemotherapy-induced nausea and vomiting among cancer patients receiving highly emetogenic AC chemotherapy, complete protection (CP) is achieved in less than 60% of patients while nearly 50% still experience nausea. Does determination of genetic constitution enable selection of the most effective antiemetic prophylaxis?

*Added value of this study:* By applying pharmacogenomic study, homozygous TT of rs1176744 genotype of HTR3B and homozygous GG of rs3821313 genotype in TACR1 were found to have better CP rates when olanzapine is being combined with aprepitant/ondansetron/dexamethasone. Digenic interaction analysis further reveals significant interaction between these genes. Olanzapine-containing regimen yielded the highest CP rates among patients who were both homozygote T of rs1176744 and homozygote G of rs3821313. Similar findings were also observed for patients who were homozygote G in both rs1176722 of HRB3A and rs3821313 of TACR1.

*Implications of all the available evidence:* Among patients who received highly emetogenic AC chemotherapy, assessment of patients’ genetic constitution can enable appropriate selection of the most optimal antiemetic prophylaxis.

## Introduction

Chemotherapy is an important component of anti-neoplastic strategy for cancer patients. However, chemotherapy-induced nausea and vomiting (CINV) is a common and distressing side effect experienced by many patients, affecting their quality of life.^1^ CINV may become so severe that treatment has to be modified or even discontinued, leading to suboptimal outcomes. The primary goal of prophylactic antiemetic treatment is to prevent CINV. International guidelines from esteemed associations, such as the National Comprehensive Cancer Network (NCCN), the American Society of Clinical Oncology (ASCO), the European Society for Medical Oncology (ESMO), and the Multinational Association of Supportive Care in Cancer (MASCC) have ranked the emetogenic risk for commonly used chemotherapy agents, whereby prophylactic antiemetics could be administered accordingly.^1–4^ For instance, the commonly administered anthracycline-cyclophosphamide (AC) regimen as (neo)adjuvant therapy in breast cancer patients has been categorized as highly emetogenic chemotherapy (HEC), with more than 90% risk of emesis, and prophylactic use of a triple-drug regimen of antagonists against receptors of 5-hydroxytryptamine type 3 (5HT3 receptor, type 3 serotonin receptor) and neurokinin-1 (NK1) and dexamethasone, is recommended.^2,4,5^ In addition, olanzapine, which is believed to achieve antiemetic control by targeting multiple pathways, has also been recommended as an additional agent in the management armamentarium.^6^ However, studies assessing the role of adding olanzapine have yielded conflicting results.^7–10,11^ This could be due to the fact that such antiemetic regimens are prescribed on an empirical basis to all patients. Complete control of CINV, in particular, that related to symptoms of nausea, is still lacking in a significant proportion of patients. At the same time, it has been well-reported that olanzapine, especially when given at a commonly adopted dose of 10 mg, commonly causes side effects of somnolence and higher tendency of non-neutropenic fever.^7–10,11^

Previous genetic studies on emesis have only focused on the genetic predisposition for CINV or postoperative nausea and vomiting.^12–17^ Few studies have evaluated the genetic determinants that could affect the variability in response to antiemetic regimens for the control of CINV. A number of genes coding for subunits of receptors of 5- hydroxytryptamine (serotonin) have been studied, including HTR3A and HTR3B.^18,19^ In addition, TACR1 gene, which codes for the neurokinin 1 receptor (also known as the tachykinin receptor 1) has also been studied.^16,20^ To date, no study has been performed comparing efficacy among different prophylactic CINV regimens in patients with different genotypes in pharmacogenetic receptor genes. Only when multiple regimens are compared could a potential more responsive prophylactic regimen be identified for patients with different genetic makeup. This will be a landmark finding for future practice of personalized medicine in this area. The results from our current study support the use of prior pharmacogenetic assessment in these three genes in the identification of appropriate prophylactic antiemetic regimens for a particular patient, a finding that has not been reported before.

In this study, we postulated that determination of these three genes may predict the response to a particular antiemetic regimen for an individual patient, thereby enabling personalized selection of optimal antiemetic prophylaxis.

## Methods

### Study design and participants

Patients in the current pharmacogenomic study participated in two previously reported prospective antiemetic studies for CINV, which were registered at ClinicalTrials.gov (Identifier: NCT03386617 and NCT03079219, respectively).^11,21^ Both studies enrolled similar patient population; that is, Chinese female breast cancer patients who were chemotherapy-naive and planning to receive (neo)adjuvant doxorubicin 60 mg/m2 and cyclophosphamide 600 mg/m2 (AC) on day 1 of a 2- or 3-week chemotherapy cycle. Study design and efficacy assessment could be obtained from previous reports.^11–21^ The antiemetic regimens are listed in Table S1. The first study was conducted between 2017 and 2018 and randomized patients to aprepitant/ondansetron/dexamethasone (for the purpose of this report, this is labeled as ‘regimen A’) or to aprepitant/ondansetron/dexamethasone with olanzapine (‘regimen B’).^11^ The second study was a prospective single arm study conducted in 2018-2019 which evaluated netupitant/palonosetron/dexamethasone (‘regimen C’, with netupitant and palonosetron combined in a capsule known as NEPA).^21^

### Assessment of chemotherapy-induced nausea and vomiting

Symptoms of CINV were captured from patients’ diaries and questionnaires during the first cycle of AC. In addition to symptoms of vomiting and the use of rescue antiemetic medications, each patient indicated her symptoms of nausea on a visual analogue scale (VAS) which ranged from 0 to 100 mm.

Nausea was the key issue to be addressed. As a result, two key endpoints were addressed, namely ‘complete protection’ (CP) and ‘no nausea’ (NN). CP was defined as no vomiting, no use of rescue therapy and ‘no significant nausea’ (NSN) during the study periods. NSN was defined as a nausea VAS of <25 mm, while NN was defined as a nausea VAS of <5 mm. The study period included acute phase (0-24 hours after initiation of AC), delayed phase (24-120 hours) and overall phase (0-120 hours) in the first cycle of AC.

### Laboratory methods for genotyping

Ten milliliters of peripheral blood was obtained from consented patients. Genomic DNAs were extracted from peripheral blood using commercial kits. PCR was carried out under standard conditions in 96- or 384-well format. Three candidate functional SNPs of three receptors genes related to 5HT3 and NK1 receptors were determined to be primary targets: HTR3A, HTR3B and TACR1 (Table 1). Genotyping for single nucleotide polymorphisms (SNPs) in the candidate genes was carried out by one of the established protocols, melting curve analysis and/or PCR-Sanger sequencing. Both positive and negative control samples were included together with replicated samples representing at least 5% of the original sample set. Any genotype data showing departure from Hardy-Weinberg equilibrium were re-genotyped by a different protocol.

**Table 1.**
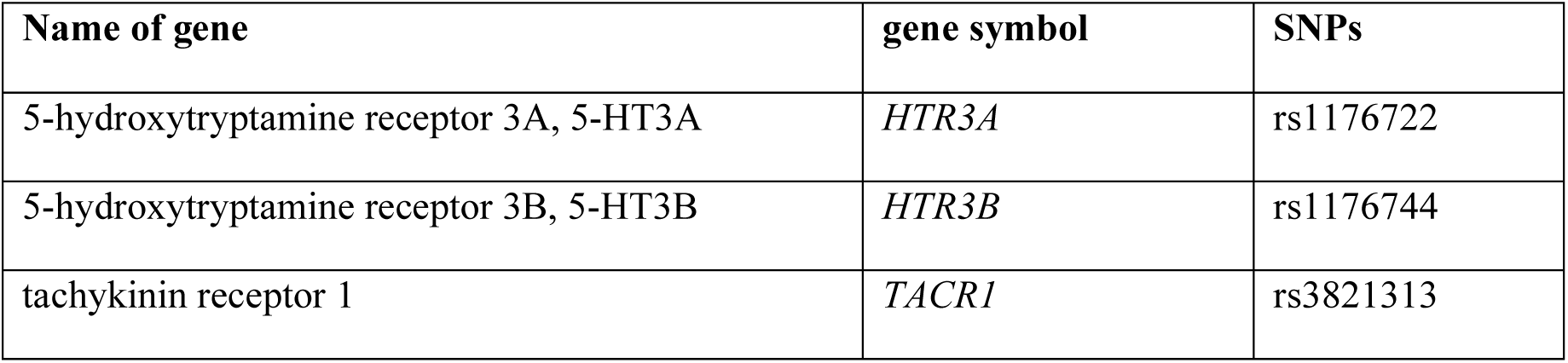
List of Genes and SNPs studies for association with anti-emetic efficacy.

The PCR-melting genotyping methodology was published in previous studies.^22,23^ Briefly, PCR reactions were carried out in a total volume of 15 µl containing 10 ng of DNA, 10 mM Tris–HCl buffer (pH 8.3), 1.5 mM MgCl2, 200 mM of each deoxynucleotide triphosphate, 50 ng of primers with different 5’ tails in the presence of Taq DNA polymerase (Roche Molecular Biochemicals). After PCR, the genotypes of samples were revealed by their melting temperature in the presence of a fluorescent DNA binding dye such as SYBR Green. DNA samples of known genotypes were included as a positive control in each batch of 96-well plates. Results were also confirmed by Sanger sequencing of PCR products.

### Statistical analysis

For genetic association analysis, each treatment regimen, patients’ response (i.e. CP or not, as well as ‘NN’ or not) was used to classify patients into 2 categorical groups. Thereafter, their genotypes of the genetic polymorphisms were compared between these 2 groups in a 2 x 3 table. A statistically significant result indicates that the genotype of that gene polymorphism determines the response or efficacy of that antiemetic regimen. Besides, the CP rate and NN rate (i.e. the proportion of patients achieving CP and NN respectively) were compared across various regimens for patients with given genotypes in order to identify the most effective treatment regimen with the corresponding genotypes. The association between CP, NN and various genotypes are reported as odd ratios (OR), with 95% confidence intervals (CIs) and p-value. The multivariate analysis, adjusting for the clinical factors, was performed. A two-sided P value < 0.05 was considered statistically significant. Statistical analysis was performed based on SAS version 9.4 (SAS Institute, Cary, NC).

### Ethics statement

Both antiemetic studies were approved by the Joint CUHK-NTEC Institutional Review Board of the Chinese University of Hong Kong and Hong Kong Hospital Authority; apart from the main study consent, patients were invited and signed a separate consent for the current study.^20,21^ The second study^21^ was also conducted in another cancer center with ethics approval from the Kowloon West Cluster Research Ethics Committee of the Hong Kong Hospital Authority; these patients (five in total) were not involved in the present pharmacogenomic study.

## Results

### Patient characteristics

A total of 180 patients took part in the two clinical studies and received Regimen A, B or C antiemetic prophylaxis. One hundred and twenty-nine patients were consented for the present pharmacogenomic studies: 45 underwent regimen A, 42 received regimen B and 42 had regimen C. The background characteristics of these patients are shown in Table S2. The proportion of patients in regimen A vs B vs C who achieved CP after AC were 38%, 57% and 55% respectively, while that for NN were 33%, 55% and 52% respectively. Overall, the efficacy of those best prophylactic regimens was around 50% with regimen B (Olanzapine-containing 4-drug regimen) and regimen C (netupitant-containing 3-drug regimen).

### Association of SNPs genotypes with antiemetic efficacy

#### Genetic association study of antiemetic efficacy in terms of Complete Protection (CP)

The most robust pharmacogenetic association was found in patients given regimen B (olanzapine-containing 4-drug regimen). Both HTR3B and TACR1 genotypes influence the prophylactic efficacy of this regimen.

Table 2 shows the genetic association analysis between various SNPs of HTR3B genotype and treatment response among patients given regimen B. For the overall phase, the T allele (encoding for Tyrosine at codon 129 of HTR3B) is the common allele of rs1176744 genotypes in HTR3B; it is also referred to as the A allele (on the complimentary sense strand) in dbSNP. This SNP is a nonsynonymous mutation; it is also known as Tyr129Ser (p.Y129S) where the tyrosine of codon 129 is substituted by serine by this T to G nucleotide change. Our results showed that homozygote patients with tyrosine of codon 129 (p.129Tyr/Tyr or TT) were more likely to achieve CP against CINV when prophylactically given the 4-drug regimen with olanzapine. Among the 29 homozygous p.129Tyr (p.129Tyr/Tyr) patients, 69% achieved CP after treatment with regimen B, while only 31% (4 out of 13) of the heterozygote GT patients (p.129Ser/Tyr) achieved CP (Odds ratio [OR]: 0.200, 95% confidence interval [CI]: 0.049-0.825; p= 0.0260).

**Table 2.** Association between HTR3B genotypes and treatment efficacy of regimen B (the olanzapine-containing 4-drug regimen)

| rs1176744 genotypes in HTR3B | GT<br>No. (%) | TT<br>No. (%) | p-value* | OR<br>(95% CI, p-value**) |
| --- | --- | --- | --- | --- |
| CP in overall phase (0-120 hr) |  |  |  |  |
| Patients did not have complete protection | 9 (70) | 9 (31) | 0.0202 | 0.200 (0.049-0.825, 0.0260) |
| Patients achieved complete protection | 4 (30) | 20 (69) |  |  |
| CP in acute phase (0-24 hr) |  |  |  |  |
| Patients did not have complete protection | 7 (54) | 6 (21) | 0.0319 | 0.224 (0.054-0.919, 0.0377) |
| Patients achieved complete protection | 6 (46) | 23 (79) |  |  |
| CP in delayed phase (24-120 hr) |  |  |  |  |
| Patients did not have complete protection | 8 (62) | 7 (24) | 0.0204 | 0.199 (0.049-0.810, 0.0242) |
| Patients achieved complete protection | 5 (38) | 22 (76) |  |  |

CP- Complete Protection
OR- Odds Ratio; 95% CI- 95% Confidence Interval
p-value\* from chi-square test or fisher-exact test
p-value\*\* from logistic regression

When the assessment period is subdivided into acute and delay phases, similar associations were found for rs1176744. During the delay phase, homozygote patients of Tyr at codon 129 (p.129Tyr/Tyr) of the HTR3B gene were more likely to achieve CP: 22 out of 29 homozygotes (76%) vs 5 out of 13 heterozygotes (38%) (OR: 0.199, 95% CI: 0.049-0.810; p= 0.0242).

An association was also found between rs3821313 in the TACR1 gene and CP rates for the acute phase among patients in regimen B (Table 3). As AA genotype is rare (∼5%), this genotype is combined with the heterozygote subjects for analysis; in other words, the results of the common allele (G) are being analyzed in a recessive mode. Homozygote G patients (GG genotype) were more likely to achieve CP during the acute phase (OR: 0.128, 95% CI: 0.024-0.687; p= 0.0165).

**Table 3.** TACR1 genotypes and treatment efficacy of regimen B (the olanzapine-containing 4-drug regimen). (a) Distribution of genotypes and (b) Association between TACR1 genotypes and treatment efficacy of regimen B.

| rs3821313 genotypes in TACR1 | AA<br>No. (%) | GA<br>No. (%) | GG<br>No. (%) | p-value* |
| --- | --- | --- | --- | --- |
| CP in overall phase (0-120 hr) |  |  |  |  |
| Patients did not have CP | 1 (25) | 11 (58) | 6 (32) | 0.1957 |
| Patients achieved CP | 3 (75) | 8 (42) | 13 (68) |  |
| CP in acute phase (0-24 hr) |  |  |  |  |
| Patients did not have CP | 1 (25) | 10 (53) | 2 (11) | 0.0187 |
| Patients achieved CP | 3 (75) | 9 (47) | 17 (89) |  |
| CP in delayed phase (24-120 hr) |  |  |  |  |
| Patients did not have CP | 0 (0) | 10 (53) | 5 (26) | 0.0699 |
| Patients achieved CP | 4 (100) | 9 (47) | 14 (74) |  |

**b)**
| rs3821313 genotypes in TACR1 | AA or GA | GG | p-value* | OR<br>(95% CI, p-value**) |
| --- | --- | --- | --- | --- |
| CP in overall phase (0-120 hr) |  |  |  |  |
| Patients did not have CP | 12 (52) | 6 (32) | 0.1795 | 0.423 (0.119-1.502, p=0.1833) |
| Patients achieved CP | 11 (48) | 13 (68) |  |  |
| CP in acute phase (0-24 hr) |  |  |  |  |
| Patients did not have CP | 11 (48) | 2 (11) | 0.0093 | 0.128 (0.024-0.687, p= 0.0165) |
| Patients achieved CP | 12 (52) | 17 (89) |  |  |
| CP in delayed phase (24-120 hr) |  |  |  |  |
| Patients did not have CP | 10 (43) | 5 (26) | 0.2479 | 0.464 (0.125-1.725, p= 0.2520) |
| Patients achieved CP | 13 (57) | 14 (74) |  |  |

CP- Complete Protection
OR- Odds Ratio; 95% CI- 95% Confidence Interval
p-value\* from chi-square test or fisher-exact test
p-value\*\* from logistic regression

#### Genetic association study of antiemetic efficacy in terms of no nausea (NN)

Only one significant association was found in patients who received regimen A (3-drug regimen with aprepitant, ondansetron and dexamethasone) with NN (Table 4). As homozygous AA of rs1176722 genotypes in HTR3A is rare (<5%), this genotype is combined with the heterozygote subjects for analysis. Homozygote patients with the G allele of rs1176722 in the HTR3A gene were more likely to have nausea during overall phase after chemotherapy (19 out of 23 GG homozygotes experienced nausea, 83%), compared to only 50% of the non-GG patients (OR: 0.211, 95% CI: 0.054-0.824; p=0.0252). Similar trends for CP rates were being observed during the acute and delayed phases.

**Table 4.** HTR3A genotypes and treatment efficacy (in terms of ‘No Nausea (NN)’ of regimen A (Aprepitant/ondansetron/dexamethasone 3-drug regimen). (a) Distribution of genotypes (%). (b) Association between HTR3A genotypes and treatment efficacy of regimen A.

| rs1176722 genotypes in HTR3A | AA | GA | GG | p-value* |
| --- | --- | --- | --- | --- |
| **NN in overall phase (0-120 hr)** |  |  |  |  |
| Patients did not have NN | 0 (0) | 11 (52) | 19 (83) | 0.0376 |
| Patients achieved NN | 1 (100) | 10 (48) | 4 (17) |  |
| **NN in acute phase (0-24 hr)** |  |  |  |  |
| Patients did not have NN | 0 (0) | 7 (33) | 13 (57) | 0.2010 |
| Patients achieved NN | 1 (100) | 14 (67) | 10 (43) |  |
| **NN in delayed phase (24-120 hr)** |  |  |  |  |
| Patients did not have NN | 0 (0) | 11 (52) | 18 (78) | 0.0796 |
| Patients achieved NN | 1 (100) | 10 (48) | 5 (22) |  |

**(b)**
| rs1176722 genotypes in HTR3A | AA or GA | GG | p-value* | OR<br>(95% CI, p-value**) |
| --- | --- | --- | --- | --- |
| NN in overall phase (0-120 hr) |  |  |  |  |
| Patients who experienced nausea | 11 (50) | 19 (83) | 0.0204 | 0.211<br>(0.054-0.824, p=0.0252) |
| Patients achieved NN | 11 (50) | 4 (17) |  |  |
| NN in acute phase (0-24 hr) |  |  |  |  |
| Patients who experienced nausea | 7 (32) | 13 (57) | 0.0955 | 0.359<br>(0.106-1.214, p= 0.0994) |
| Patients achieved NN | 15 (68) | 10 (43) |  |  |
| NN in delayed phase (24-120 hr) |  |  |  |  |
| Patients who experienced nausea | 11 (50) | 18 (78) | 0.0477 | 0.278<br>(0.076-1.015, p= 0.0528) |
| Patients achieved NN | 11 (50) | 5 (22) |  |  |

NN= No Nausea
OR- Odds Ratio; 95% CI- 95% Confidence Interval
\*p-value from chi-square test or fisher-exact test
\*\* p-value from logistic regression

For instance, in the delayed phase, 78% of homozygous GG experienced nausea; this contrasted with other patients, where 50% experienced nausea (OR: 0.278, 95% CI: 0.076-1.015, p= 0.0528).

#### Gene-gene interaction in the prophylactic treatment of CINV - HTR3B and TACR1 digenic genotype effects

The effects of a combination of genotypes in 2 SNPs were analyzed. Specifically, only those SNPs with a significant association with treatment efficacy were analyzed. CP in the overall phase was used as the treatment outcome.

Interaction between rs1176744 (HTR3B) and rs3821313 (TACR1) was found to affect the treatment efficacy of prophylactic antiemetic regimens. Figure 1 shows the percentage of patients who achieved CP with the 3 prophylactic antiemetic regimens. The G allele of rs1176744 and A allele of rs3821313 were minor alleles of low allelic frequencies in the population, so no patient being homozygous for both of these alleles were studied (Figure 1A). On the other hand, many patients were both homozygote T of rs1176744 and homozygote G of rs3821313 and their treatment efficacy toward the 3 regimens is shown in Figure 1I. There was a striking difference among the 3 regimens in terms of percentages of patients achieving CP with this genotypic combination in the HTR3B and TACR1 genes. This group of patients had the higher CP rate by regimen B (crosshatch bar in Figure 1I), with 10 out of 12 (83.3%) patients achieving CP in the overall phase. On the other hand, only 29% (7 out of 24) of patients given regimen A (stripe bar in Figure 1I) experienced CP while 50% (7 out of 14) of patients achieved CP with regimen C (p= 0.0089).

**Figure 1.**
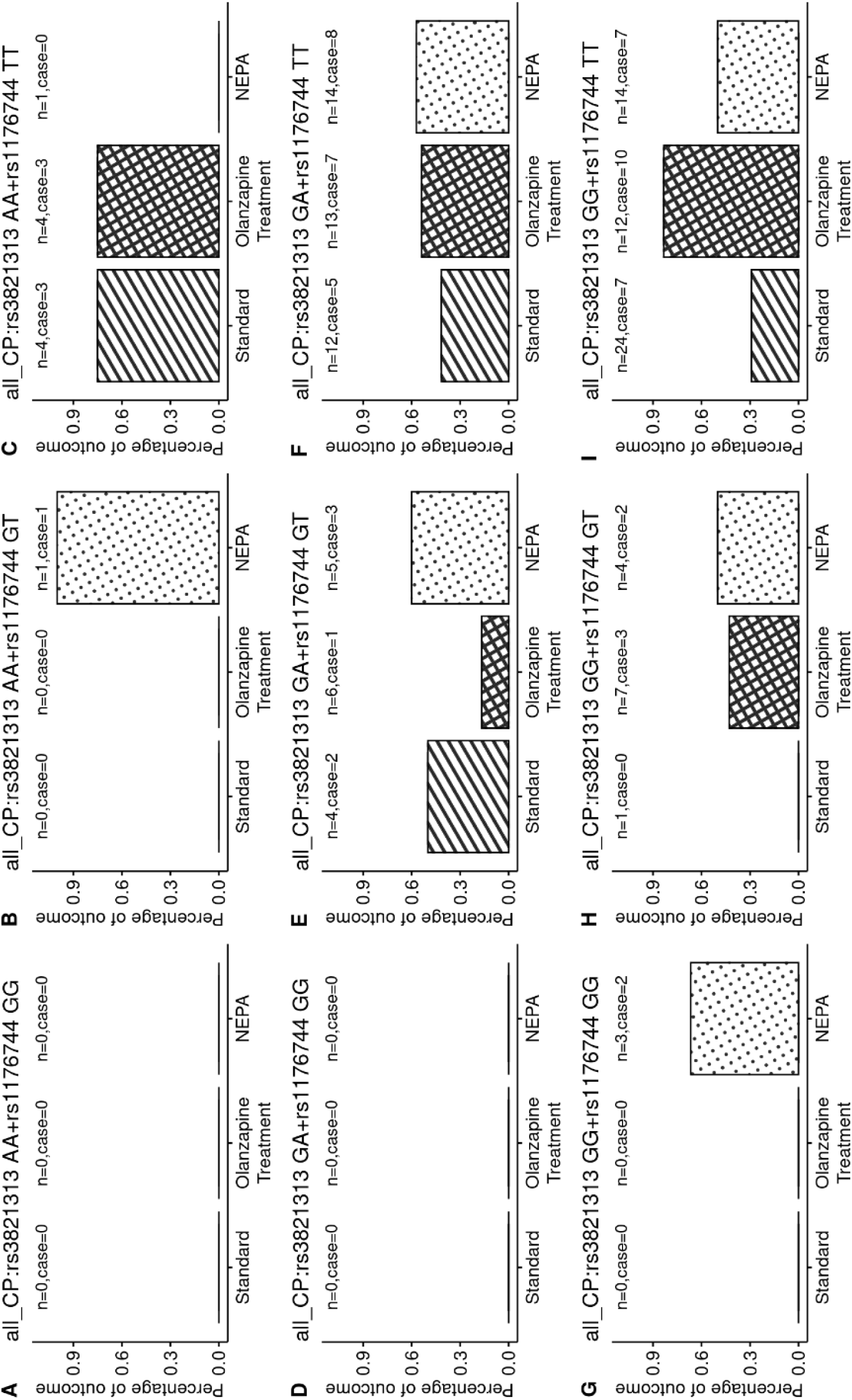
Interaction between rs1176744 (HTR3B) and rs3821313 (TACR1) determines response to various prophylactic antiemetic treatment regimens in chemotherapy patients. (Standard: regimen A- aprepitant/ondansetron/dexamethasone, Olanzapine: regimen B- olanzapine/aprepitant/ondansetron/dexamethasone and NEPA: regimen C- netupitant/ondansetron/dexamethasone). The numbers of patients in each group (n=) are shown above each bar together with numbers that had complete protection which is labelled as case (case=). Y axis shows the percentage of patients who achieved complete protection for each regimen.

As the minor alleles of both SNPs are uncommon in the population, the digenic interaction was analyzed by reducing the 3×3 genotype combinations into 2×2 genotype combinations by combining the homozygotes of minor alleles with heterozygotes into one class, as shown in Figure 2 (upper left inset). In terms of biology, this assumes that the major alleles act in a recessive manner.^23^ The results after such rearrangement confirmed that patients who were homozygotes of the common alleles in both SNPs (rs1176744 and rs3821313) had the highest CP rate with regimen B (i.e. adding olanzapine to the 3-drug regimen) (crosshatch bar in Figure 2D) than regimen A (stripe bar in Figure 2D), 83% vs 29% respectively, a difference confirmed to be statistically significant (Fisher exact test, p= 0.0027).

**Figure 2.**
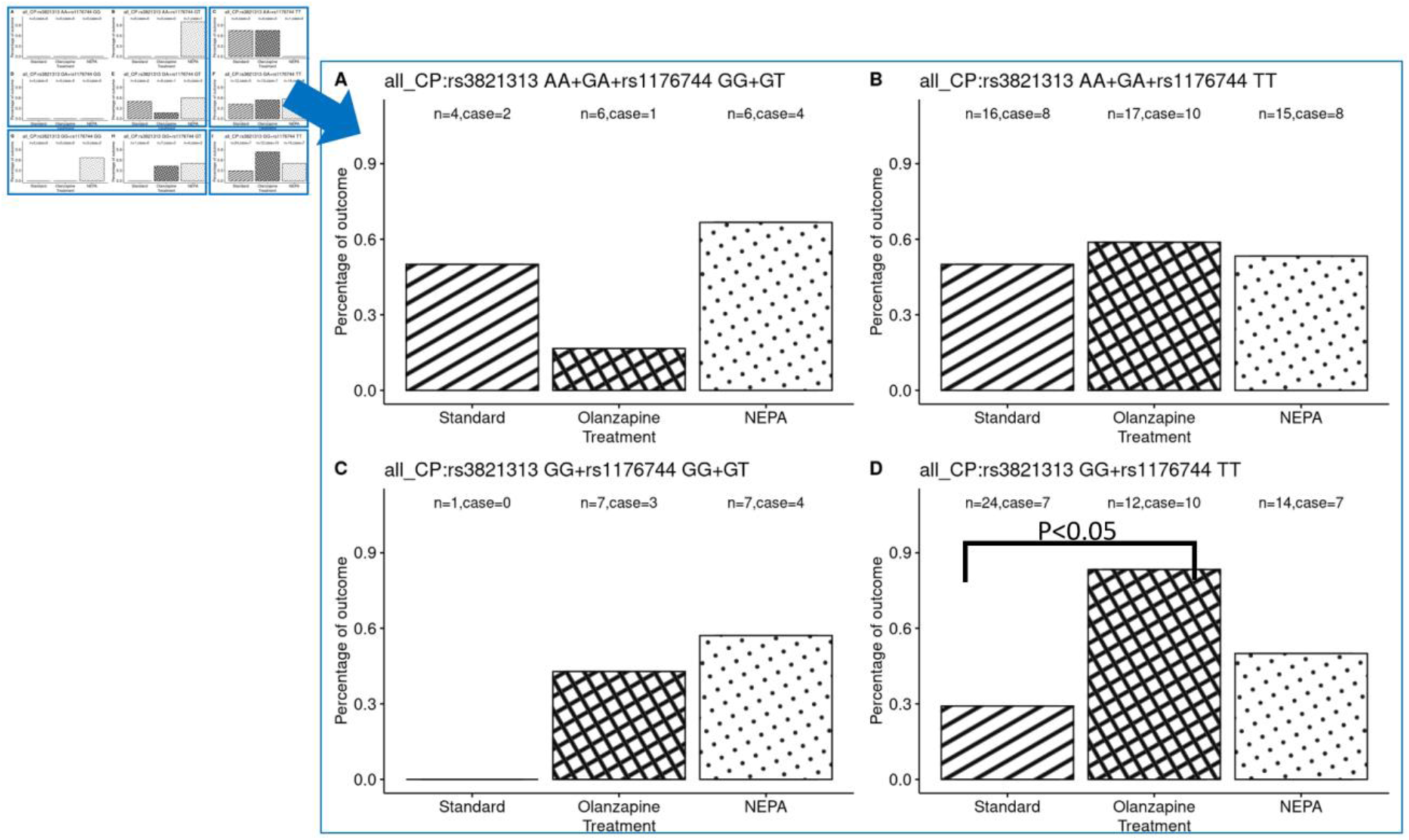
Using a recessive mode to analyze the interaction between rs1176744 (HTR3B) and rs3821313 (TACR1) and response to various prophylactic antiemetic regimens in chemotherapy patients. Y axis shows the percentage of patients who achieved complete protection for each regimen. Assuming the gene-gene interaction occurs in the homozygotes of both major alleles in a recessive mode, the 3 x 3 combination of genotypes (left upper) are combined as shown into 4 groups of digenic genotypes as shown in the right lower larger graph. (Standard: regimen A-aprepitant/ondansetron/dexamethasone, Olanzapine: regimen B-olanzapine/aprepitant/ondansetron/dexamethasone and NEPA: regimen C-netupitant/ondansetron/dexamethasone). The numbers of patients in each group (n=) are shown above each bar together with numbers that had complete protection which is labelled as case (case=). In Figure 2D, the P value of statistical comparison between regimen A (Standard) and regimen B (olanzapine-containing) is shown.

On the other hand, patients with combinations of genotypes rs1176744 (GG or GT) and rs3821313 (AA or GA) had the highest CP rate (67%) with regimen C (NEPA-containing regimen) (dotted bar in Figure 2A), as compared to regimen A (CP rate = 50%) or regimen B (CP rate= 17%), though not reaching statistical significance (p= 0.2089).

#### Gene-gene interaction in the prophylactic treatment of CINV - HTR3A and TACR1 digenic genotype effects

Interaction between rs1176722 (HTR3A) and rs3821313 (TACR1) also affects the treatment efficacy of prophylactic antiemetic regimens. A similar approach was used to combine genotypes of these 2 SNPs into a 2×2 genotype combination figure (Figure 3). Figure 3D showed the treatment efficacy in terms of CP rates in the overall phase using the three prophylactic antiemetic regimens. Patients who were homozygote for G alleles in both rs1176722 and rs3821313 showed the poorest response to regimen A with CP rate of 17% (2 out of 12 had CP, stripe bar in Figure 3D), while patients given regimen B had the highest CP rate of 70% (7 out of 10 had CP) (regimen A vs B, p= 0.0159). Regimen C was numerically superior to regimen A, with CP rates of 46% vs 17% respectively, though not reaching statistical significance (p= 0.1047).

**Figure 3.**
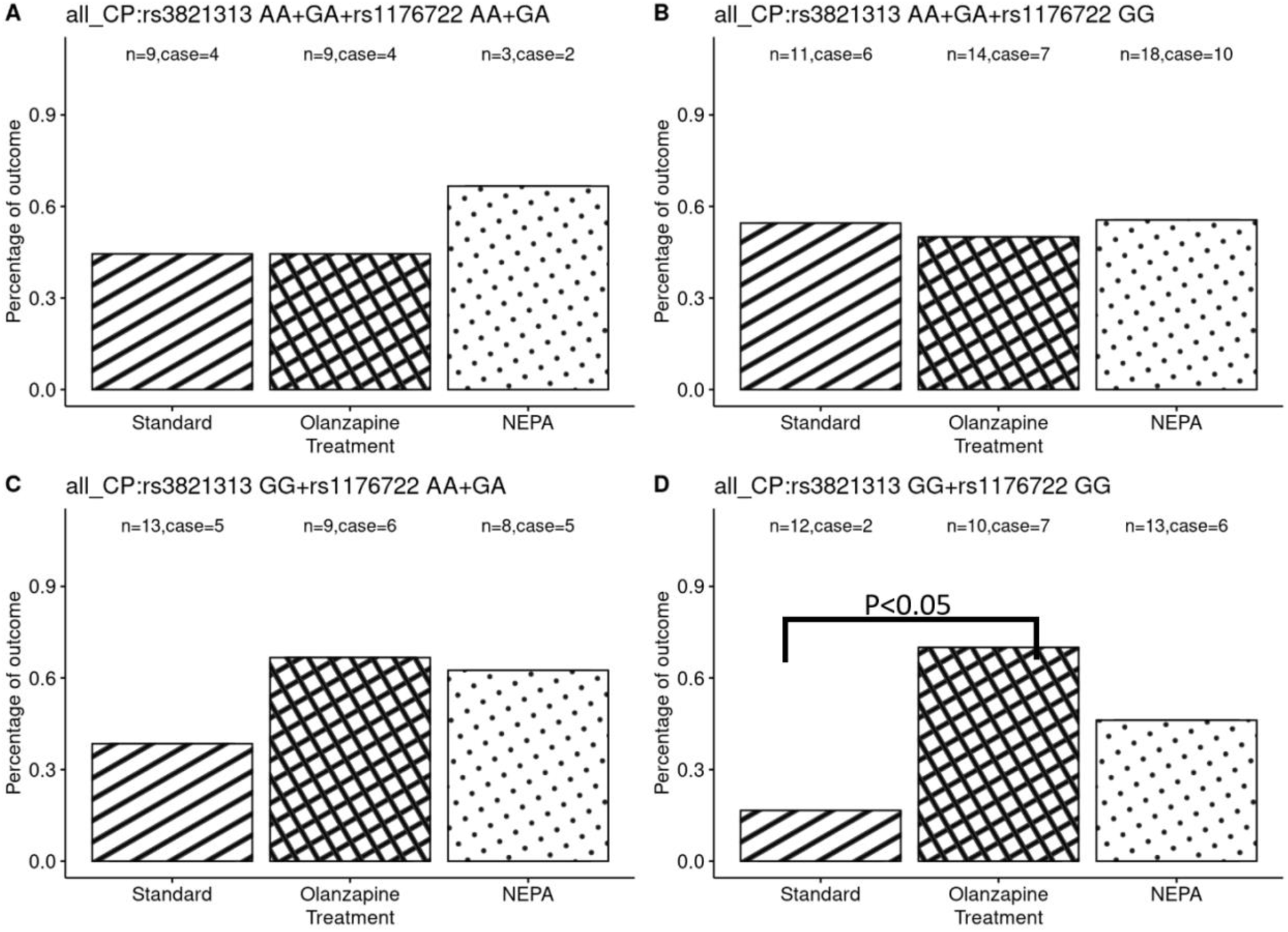
Using a recessive mode to analyze the interaction between rs1176722 (HTR3A) and rs3821313 (TACR1) and response to various prophylactic antiemetic regimens in chemotherapy patients. Assuming the gene-gene interaction occurs in the homozygotes of both major alleles in a recessive mode, the 3 x 3 combination of genotypes of these 2 SNPs are combined as shown into 4 groups of digenic genotypes as here. (Standard: regimen A-aprepitant/ondansetron/dexamethasone, Olanzapine: regimen B-olanzapine/aprepitant/ondansetron/dexamethasone and NEPA: regimen C-netupitant/ondansetron/dexamethasone). The numbers of patients in each group (n=) are shown above each bar together with numbers that had complete protection which is labelled as case (case=). In Figure 3D, the P value of statistical comparison between regimen A (Standard) and regimen B (Olanzapine-containing) is shown.

On the other hand, patients with combinations of genotypes rs1176722 (AA or GA) and rs3821313 (AA or GA), as shown in Figure 3A, experienced a relatively higher CP rate of 67% with regimen C (2 out of 3 patients, dotted bar in Figure 3A), while that of regimen A and B were similar at 46% (p= 0.7753).

### Multivariate analysis of genetic and clinical risk factors for CINV

After being adjusted for clinical factors, logistic regression revealed that CP rates in all phases among patients who underwent regimen B were significantly higher with rs1176744 homozygous TT in HTR3B. The ORs (95% CI, p-value) for overall, acute and delayed phases for this genotypic were respectively 0.200 (0.049-0.825, p= 0.0260), 0.070 (0.008-0.658, p= 0.0200), and 0.199 (0.049-0.810, p= 0.0242).

In addition, rs3821313 homozygous GG in TACR1 was associated with significantly higher CP rate in the acute phase (OR: 0.045, 95% CI: 0.004-0.492, p= 0.0110).

## Discussion

International guidelines have recommended 3-drug antiemetic prophylaxis regimen of 5HT3-and NK1-receptor antagonist in combination with dexamethasone as an option for cancer patients to prevent CINV, mainly based on their efficacy in control acute phase CINV. Although a number of randomized controlled trials have demonstrated that adding olanzapine to triplet antiemetic regimen such as aprepitant/ondansetron/dexamethasone could improve CINV control among cancer patients who received HEC regimen,^6,7,24^ delayed phase CINV, especially related to nausea, remains to be a distressing symptom in a significant proportion of patients.^11,21^ This is reflected by our findings that with the relatively older 3-drug regimen (A) of aprepitant/ondansetron/dexamethasone, the CP and NN rates were only 38% and 33% respectively. Even with more contemporary antiemetic prophylaxis, over 40% of patients given olanzapine/aprepitant/ondansetron/dexamethasone (regimen B) did not achieve CP and NN respectively. Similarly, using a combination of netupitant/palonosetron/ dexamethasone (regimen C), a second generation NK1- and 5HT3- receptor antagonists, the CP and NN rates were only 55% CP rate and 53% NSN rate, respectively. Further, olanzapine has been well-associated with the adverse effects of somnolence and fever.^7–11,20^

Previous genetic studies in relation to CINV have assessed genetic polymorphisms of transporters of central nervous system, drug metabolisms and target receptors of antiemetic agents in association with risk of CINV. The genetic polymorphisms assessed have included ATP binding cassette subfamilies ABCB1 and ABCB2, and target receptors related to 5HT3 and NK1, including HTR3A/HTR3B/HTR3C/HTR3D and TACR1 respectively.^12–20^ These studies have either failed to identify relevant genetic polymorphisms or have been inconsistent in their findings. To further decipher the relevance of these polymorphisms in CINV, a meta-analysis was conducted by Eliasen et al^12^ and involved 20 studies with over two thousand patients. Eight polymorphisms in five candidate genes were analyzed. Only HTR3C and ABCB1 polymorphisms were identified to be associated with acute CINV. Such negative findings could be due to inclusion of a large range of genetic polymorphisms which were evaluated in a heterogeneous group of patient populations who received different chemotherapy of varied emetogenic potential.

Pharmacogenomic analysis, by assessing genetic polymorphisms in relation to effectiveness of specific prophylactic antiemetic regimens, have not been well-studied. While the initial report of suggested that ABCB1 and ABCB2 genetic polymorphisms were associated with acute phase CINV development,^25^ their follow-up study was unable to confirm such earlier findings.^15^ Using a uniform antiemetic prophylaxis of granisetron/dexamethasone/aprepitant, the authors also assessed target receptors related to 5HT3 and NK1, namely HTR3B/HTR3C/HTR3D and TACR1 respectively. While there were no significant findings with genes associated with 5HT3 receptors, TACR1 was suggested to be a potential genetic risk factor for delayed CINV.^16,20^

Our study has some limitations, mainly related to small patient number of one gender receiving one chemotherapeutic regimen only. None-the-less, the current pharmacogenomic study has several strengths and is unique in a number of ways. A homogenous patient population of Chinese ethincity with breast cancer who were chemo-naïve and receiving AC was studied. Having involved patients from two prospective antiemetic studies means that the emetic events were captured accurately, further strengthening the findings of this study. As opposed to older antiemetic prophylaxis that involved historical first generation 5HT3- receptor antagonists in combination with dexamethasone, and the first-generation NK1- antagonist aprepitant with ondansetron/dexamethasone, which we have previously reported to be inferior regimens, this study also included patients who were treated with more contemporary prophylactic regimens. The latter included the 4-drug regimen of olanzapine/aprepitant/ondansetron/dexamethasone and the 3-drug regimen of netupitant/palonosetron/dexamethasone, which have been reported to be more effective.^26,27^ Moreover, the present efficacy analysis focused on important clinical issues of chemotherapy-induced nausea, a symptom that remains to be a challenge in the prevention of CINV. Therefore, two relevant clinical endpoints were assessed instead of using the commonly assessed ‘complete response’ criteria (defined as no vomiting and no use of rescue therapy): nausea per se as well as ‘complete protection’ (which encompasses the assessment of vomiting, use of rescue therapy and nausea) were assessed.

Several findings were clinically relevant in this study. Firstly, amongst patients receiving regimen B (the 4-drug regimen of olanzapine/aprepitant/ondansetron/dexamethasone) homozygous TT of rs1176744 in the HTR3B gene achieved a significantly higher CP rate during the acute, delayed and overall phases of CINV, suggesting that TT homozygotes are best protected from CINV with regimen B. Secondly, for patients receiving the regimen B, GG homozygotes of rs3821313 in the TACR1 gene also achieved a significantly higher rate of CP in the acute phase. These pharmacogenomic findings remain to be significant upon multivariate analysis with other clinical risk factors of CINV. On the other hand, among patients given Regimen A (the 3-drug regimen of aprepitant/ondansetron/dexamethasone), GG homozygotes of rs1176722 in the HTR3A gene had significantly higher rates of nausea during the overall phase (83%) and delayed phase (78%) after chemotherapy, suggesting that this genotype should not be given regimen A alone. Finally, the current genetic findings did not affect the efficacy of Regimen C (the 3-drug regimen of netupitant/ondansetron/dexamethasone), where the CP rates were 50% or above irrespective of genotypic compositions.

Further, since it is well-known in the field of animal breeding that epistasis may occur and phenotypes may also be determined by the effect of 2 genes,^23,28,29^ the genotype patterns based on co-existence of 2 or more genes, i.e. gene-gene interaction in associated with antiemetic efficacy, was assessed.^23,28,29^ Digenic genotype interaction study between rs1176744 (HTR3B) and rs3821313 (TACR1) revealed that homozygote TT of rs1176744 and homozygote GG of rs3821313 were best treated by regimen B (CP rate of 83%) while CINV was least controlled by regimen A (CP rate of 29%). Conversely, for homozygote GG in both rs1176722 and rs3821313, regimen A should be avoided since it provided the lowest CP rate at 17% while regimen B, providing a CP rate of 70%, could be recommended. Additionally, although not statistically significant, patients with combinations of genotypes rs1176722 (AA or GA) and rs3821313 (AA or GA), as well as those with combinations of genotypes rs1176744 (GG or GT) and rs3821313 (AA or GA) had the highest CP rates with regimen C. Finally, irrespective of the genotypic variation, regimen C has consistently shown to outperform regimen A in the current genotypic analysis (Figure 2 and 3). These findings suggest that among patients with genotypic assessment revealing rs1176744 (GG or GT) or rs1176722 (AA or GA) in combination with rs3821313 (AA or GA), regimen C is the preferred antiemetic prophylaxis. Moreover, in the absence of pharmacogenetics analysis, regimen C could be regarded as a relatively more acceptable antiemetic regimen. Table 5 tabulates the above suggested antiemetic prophylaxis regimens in accordance with genotypic variables.

**Table 5.**
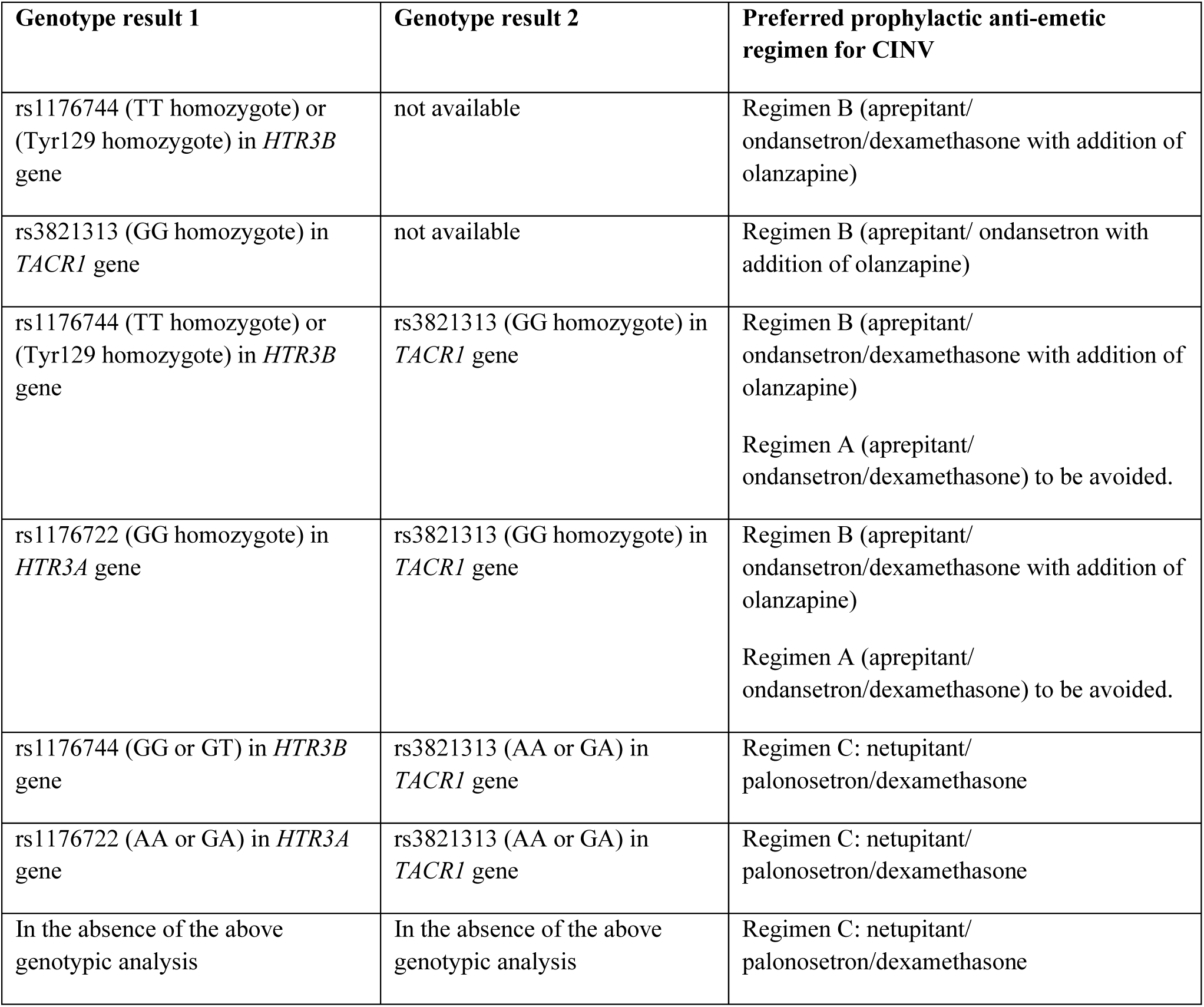
Proposed optimal antiemetic prophylaxis for individual breast cancer patients undergoing AC chemotherapy based on pharmacogenomic profiles.

We have previously reported that among patients receiving various prophylactic antiemetic regimens, those who underwent aprepitant/ondansetron/dexamethasone/olanzapine or netupitant/ palonosetron/dexamethasone had the best emetic control; however, CP and NN rates, even with these relatively better antiemetic prophylaxis, remain to be modest in the range of 55%.^26^ Based on our study, we were able to establish promising pharmacogenomic markers that can predict the efficacy of selected antiemetic regimens. Adopting the present genetic analysis has the potential to enable provision of optimal antiemetic prophylaxis to breast cancer patients planning for AC chemotherapy by increasing complete protection from CINV to up to 83%. Such judicious use of olanzapine may also help avoid unnecessary olanzapine-associated side effects for patients who would not benefit from its anti-nausea activity. However, as the current findings are limited by a relatively small patient number, additional research is needed to validate these results.

In conclusion, in addition to clinical-and treatment-related profiles, a personalized approach with the incorporation of pharmacogenomic analysis is warranted in the prevention of CINV. The current study supports the notion the pharmacogenomic analysis is feasible and could be an important element in precision medicine. In addition to symptoms of vomiting, future studies would focus on symptoms related to nausea. A hybrid combination of regimen B and C, for instance, with netupitant, palonosetron, dexamethasone and olanzapine in selected patient population based on pharmacogenetic analysis should be tested to further optimize antiemetic prophylaxis.

## Contributions

WY and NLST conceived and designed the study. FKFM, JHH, MO and NLST contributed to formal analysis. FKFM, HLY, NWHK, KTL and EP contributed to the underlying data verification. WY and FMKF had full access to all the data in the study. WY,LVL, TKHL, KTL and EP administered the project. FKFM, KTL and NLST contributed to preparation of resources and software. WY supervised this study and funding acquisition. All authors contributed to the investigation, took part in drafting, revising or critically reviewing the manuscript and approved the final manuscript for publication.

## Data sharing statement

All data relevant to the study are included in the article or uploaded as supplementary information. The data are available from the Comprehensive Cancer Trials Unit of the Department of Clinical Oncology, Chinese University of Hong Kong, but restrictions apply to the availability of these data. These data were used under permission for the current study, and so are not publicly available. Data are, however, available from the authors (WY and FM) upon reasonable request. Data will be made available for 15 years from the start of the clinical trials.

## Acknowledgement

This study was supported by an education grant from Madam Diana Hon Fun Kong Donation for Cancer Research.

